# Modern contraceptive utilization and associated factors among postpartum women, Sude District, Arsi Zone, Oromia, Ethiopia

**DOI:** 10.1101/2025.01.15.25320617

**Authors:** Mohammed Sultan Gemeda, Gemechu Dereje Feyissa, Daniel G/tsadik, Endashaw Mandefro, Hunde Lemi, Mokonnen Dereje

## Abstract

**Background:** Maternal health problems remain a major global concern since pregnancy and childbirth are the leading causes of morbidity and mortality among reproductive- age women who have recently given birth and are among the group with the highest unmet need for contraception. After a woman has given birth, she should stay for at least two years before the next pregnancy to reduce the risk of adverse maternal, pre-natal, and infant outcomes. However, postpartum modern contraceptive utilization was not studied in the study area.

**Objectives:** It aimed to assess the postpartum modern contraceptive utilization and associated factors among women who gave birth at health facilities in Sude district, Arsi Zone, Oromia, Ethiopia, 2023.

**Methods:** An institution-based cross-sectional study design was conducted from February 30 to April 30, 2023. The study participants were selected by systematic random sampling. An interviewer-administered structured questionnaire was used to collect data. After checking for its completeness, the data were entered, checked for missing values and outliers by Epi-info Version 7.2, and then exported to Statistical Package for Social Sciences (SPSS) version 25 for analysis. Data were analyzed using descriptive statistics to characterize the study participants. Binary logistic regression analysis was undertaken to identify factors associated with postpartum modern contraceptive utilization. Variables with p-value < 0.25 in bivariate analysis were considered for multiple logistic regression. Variables with a p-value < 0.05 were considered statistically significant. An adjusted odds ratio (AOR) along with 95% confidence intervals (CI) was calculated to estimate the magnitude of associations.

**Results:** Out of 419 planned, 417 participated with a response rate of 99.5%. This study revealed that 57.6% [95% CI: (52.9, 62.4)] of postpartum women utilized modern contraceptives. Among the utilizers, injectables (50.8%), implants (39.2%), and pills (10%) were used. Having > 4 children [AOR = 2.02, 95% CI: (1.09, 3.77)], ≤ 2 hours far away from home to the health facility [AOR = 2.54, 95% CI: (1.49, 4.34)], between 6 to 12 months of postpartum period [AOR = 3.41, 95% CI: (1.89, 6.15)], resumption of menses [AOR = 8.24, 95% CI: (4.64, 14.64)], para 2-4 [AOR = 3.10, 95% CI: (1.53, 6.29)], and discussing postpartum family planning with a partner [AOR = 2.96, 95% CI: (1.72, 5.09)] were significantly associated with postpartum modern contraceptive utilization.

**Conclusion:** Postpartum modern contraceptive use was low as compared to other studies. The factors associated with postpartum modern contraceptive use were: the number of children, distance from home to health facility, postpartum period, parity, and menses returning since birth, and discussing postpartum family planning with a partner. Healthcare providers should give health information about timely contraceptive usage, fortify the integration of modern contraceptive services with maternal and child health services, and encourage postpartum family planning discussions among couples.

## Introduction

Postpartum contraceptive use is defined as the avoidance of closely spaced pregnancies and unintended pregnancy during the first 12 months after delivery (1). By using a contraceptive, mothers can attain their fertility objective by providing for them to gap their pregnancies and frontier numbers of births (2). Evidence has shown that encouraging early antenatal care visits, postnatal care, institutional deliveries, and contraceptive adoption are the key elements in improving safe motherhood (3).

According to the World Health Organization (WHO), an interval of at least 2 years following a live birth is recommended for better maternal and child health outcomes before becoming pregnant again (4). Modern contraceptive plays a key role in reducing maternal and newborn morbidity and mortality by preventing unintended pregnancy and short birth intervals (5). In particular, the postpartum period is critical and is the time when many routine interventions are provided to mothers; besides, during the postpartum period, most mothers want to delay or stop the next pregnancy to reduce the risks of short-spaced, unwanted pregnancies and associated fetomaternal bad outcomes (6).

Family planning (FP) is widely acknowledged as an effective intervention for saving women’s and children’s lives and improving their health (7). It is an essential component of health care provided during the antenatal period, immediately after delivery, and during the first year postpartum (8). If a woman had only the number of pregnancies they wanted, at the intervals they wanted, maternal mortality would drop by 30% (5). Family planning (FP) can avert 3.2 million out of 5.6 million under-five deaths and 109,000 out of 155,000 (70%) of maternal deaths. However, Demographic health survey data from 57 countries indicated that, right after delivery 62%, after 6 months of amenorrhea 43% and at the end of amenorrhea 32% of women in the first year after birth have an unmet need for contraception (9).

According to the World Health Organization’s recommendation, after a woman has given birth, she should stay for at least two years before the next pregnancy to reduce the risk of adverse maternal, pre-natal, and infant outcomes (10). In Ethiopia, maternal mortality is an important public health issue;412 per 100,000 live births are dying as a result of complications related to pregnancy and childbirth (11). Closely spaced pregnancies within twelve months of delivery increase the risks of preterm birth, low birth weight, and small-for-gestational-age babies, risk of child mortality will also increase (12).

Worldwide, more than 90 percent of mothers need to hold up or evade pregnancy within the first year of the postnatal period (13). Conversely, the majority of women start sex before the first menses next to childbirth devoid of contraceptive use (14). Postpartum family planning (PPFP) utilization is inconsistent in Sub-Saharan countries, for example, less than 10% in Ethiopia, 15% in Nigeria, 20% in Tanzania, 25% in Kenya, and 40% in Zambia (15).

In Ethiopia, the risk of pregnancy among mothers who are sexually active in 12-23 months of the postnatal period is 72%, but it decreases to 64% and 37% for mothers in 6–11 and first 6 months of the postnatal period, respectively (8). Although the prevalence of postpartum mothers points out the need to utilize contraceptives, contraceptive uptakes are often not obtainable or in use by the first year of the postpartum period. The contraceptive uptake during the postpartum period in Gondar, Ethiopia, was 48.4 (16).

The Ethiopian Demographic Health Survey (EDHS) 2016 reported that 1in 5 married women had an unmet need for FP (11). Postpartum women are an important group because, even if they are breastfeeding, they may not recognize they are at risk of unintended pregnancy (17). Evidence in Ethiopia shows that almost half (47%) of all pregnancies occur within a short birth interval of less than 24 months after the previous birth (18). The current study aimed to fill this gap by assessing postpartum modern contraceptive methods utilization and identifying factors affecting their utilization in a study area. The findings of this study might be important in designing, implementing, and monitoring effective maternal and child health interventions.

## Materials and methods

### Study area and period

The study was conducted in Sude Woreda, Arsi Zone, Oromia region, Ethiopia. It is situated 96 km from Asella and 214 km from Addis Ababa, the capital city of Ethiopia. The Boundaries are Amigna district in the North, Chole district in the East, Robe district in the South, and Deksis district in the West direction. According to figures from the Central Statistical Agency (CSA) in 2015, the estimated total population of the district is 219,719 (19). The number of women attending the delivery services in the last three months in the Hospital and Health Center (HC) was 858. The health service coverage of the study area was 78% by the health center. The proportion of ANC utilization in the study area was 84%. In the Sude district, there are eight public health institutions (one public hospital and seven health centers). The health centers were called Alemgana HC, Deraba HC, Gersa Cisa HC, Halila HC, Kona Motuma HC, Kula HC, and Semar Semariyi HC. The Woreda had 27 rural and 3 urban kebeles (small administrative units). Agriculture is the main source of income for the majority of the population. The main staple food of the population was wheat and some parts barley (20). The study was conducted from February 30, 2023, to April 30, 2023.

### Study design, study population, and inclusion criteria

An institution-based cross-sectional study design was conducted. All reproductive age (15–49) women who gave birth in the last 12 months and lived in Sude district were the source population. The study population includes all post-partum women attending service delivery in Sude district public health facilities during the study period. Postpartum women who delivered a maximum of 12 months ago were included in the study whereas those who were not willing to participate, critically ill, and unable to communicate during the data collection period were excluded from the study.

### Sample size determination

The sample size was determined using a single population proportion with the statistical assumptions of 95% confidence level (Zα/2 =1.96), d = margin of error (5%), p=prevalence of postpartum modern contraceptive utilization, n = the required minimum sample size. The prevalence of postpartum family planning utilization was 54.7 % which is obtained from a study conducted in South Gondar (17). The sample size was determined as illustrated below:

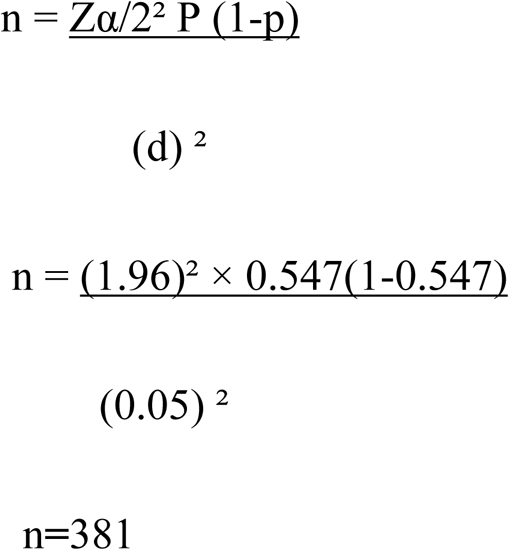

To minimize errors arising from the likelihood of non-response, 10% of the sample size was added to the estimated sample. Accordingly, the final sample size was 419.

### Sampling technique

A simple random sampling technique was employed for the selection of three health centers from seven of them and one Hospital in the District was included in this study. Then a sample frame was prepared in all selected Health Facilities to identify women who fulfill the inclusion criteria by having registered the birth date of the last child from the delivery register which was found by health workers. Based on the sample frame, the source population of each health facility was taken from several pregnant women who sought delivered services in the previous three months in the Sude District Public Health Facilities which is (858). Then, the proportional-to-size allocation technique was employed to determine the study participants from each health facility (**Fig 1).**

**Fig 1:**
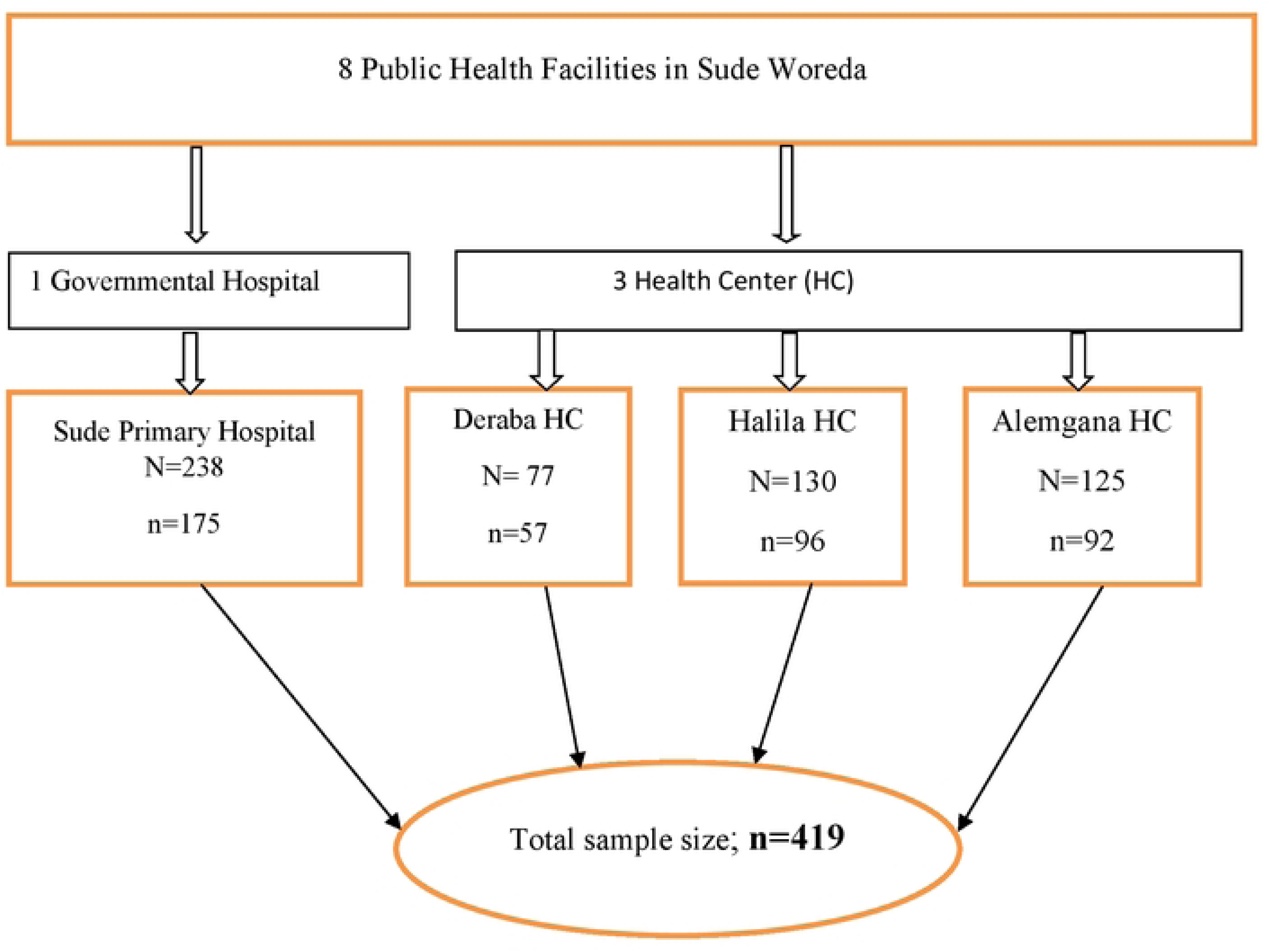
Schematic sampling procedure for assessment of postpartum modern contraceptive utilization and associated factors among women delivered in health facilities in Sude District, Arsi Zone, Oromia, Ethiopia, 2023.

Finally, reproductive-age women whose last delivery was within the past year before the study were selected using a systematic random sampling technique from the existing sampling frame in each health facility. Every third woman was interviewed after service delivery until the required sample size was obtained by taking a sample size of 419 proportionate probability sampling was used as follows j = 1, 2, k where k is the number of strata and nj is the sample size of the jth stratum Nj is population size of the jth stratum n = n1 + n2 + …+ nk is the total sample size N = N1 + N2 + …+ Nk is the total population size

### Study variables

#### Dependent variable

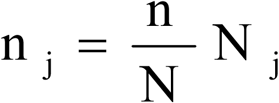

Postpartum modern contraceptive utilization

#### Independent variables

Socio-demographic characteristics such as age, marital status, educational status, religion, occupational status of the mother, husband’s educational status, and occupation. Reproductive and maternal health service use-related characteristics include parity, birth space, discussion with husband about postpartum family planning, previous history of contraceptive utilization, antenatal care follow-up, and postnatal care follow-up, places of delivery, family planning counseling, birth attendants, menses return, length of time after delivery, pregnancy plan, want to have children for the future, and resumed sexual intercourse. Knowledge on family planning such as awareness of contraceptive use after birth, source of knowledge, source of family planning, and the benefit of family planning.

### Operational definition

#### Modern contraceptives

Injectable, intrauterine devices (IUDs), contraceptive pills, implants, male and female sterilization, emergency contraception, female and male condoms, and the standard day’s method were considered modern contraceptives (21).

#### Postpartum contraceptive utilization

women were considered as utilized when the women reported that they used any type of modern contraceptive method by themselves or jointly with their husbands from data collection time to the last 12 months after birth (17). The utilization was measured by mothers’ words by yes or no options for use (Yes = 1, No = 0).

### Data collection procedures

The data was collected by using a pre-tested interview administrated semi-structured questionnaire. The questionnaire was first developed in English and translated into the local language (Afan Oromo) and then back to English by language experts to keep its consistency. The questionnaire contains sections for assessing demographics and associated factors. The questions and statements were grouped and arranged according to the particular objectives that they aimed to address the objectives. Six BSc Midwifery and two MPH who were familiar with the local language and customs were recruited as data collectors and supervisors respectively. The training was given to data collectors and supervisors for two days on data collection procedures, interview techniques, and confidentiality of the information obtained from the respondents. Finally, data collectors interviewed participants face-to-face at health facilities for 20–25 minutes.

### Data quality control

Data quality was ensured during collection, entry, and analysis. Before conducting the main study, a pretest was carried out on 21 postpartum women at Robe General Hospital, and based on the findings necessary modification was made. The principal investigator and supervisors conducted day-to-day on-site supervision during the whole period of data collection. At the end of each day, the questionnaire was reviewed and checked for completeness and accuracy by the supervisors and investigator. The data were double-entered to verify the missing values.

### Data processing and analysis

The data were coded, checked for completeness, and entered into Epi-Info-7 then exported to SPSS version 25 for data processing and analysis. Descriptive statistics were computed and presented in frequencies and percentages for categorical variables, while mean with standard deviations were reported for continuous variables. The association between the predictors and outcome variables was conducted using bivariate and multivariate logistic regression models. Bivariate logistic regression analysis was used to assess the crude relationship between independent variables and the outcome variable. Variable in the bivariate logistic regression with a p-value of ≤ 0.25 was considered for the multivariate logistic regression. The regression model was developed using enter methods. The final fitted model was assessed for multicollinearity using the variance inflation factor (VIF) and goodness of fit using the Hosmer and Lemeshow test. Adjusted odds ratios (AOR) and the corresponding 95% confidence intervals (CI) were estimated to assess the strength of the association, and statistical significance was declared at p-value < 0.05.

### Ethics approval and consent to participate

First, ethical clearance was obtained from the Institutional Review Committee of Rift Valley University. Then, letters of permission were sought from the Sude Woreda Health Office. Following approval of ethical clearance and permission to conduct the research in the selected health facilities of the Sude District, verbal informed consent was obtained from each respondent, after giving a clear explanation about the objectives and procedures of the study before the actual data collection. A right thumbprint was received as a signature for respondents who couldn’t read and write. Respondents under the age of eighteen had their consent obtained from their parents or legal guardians. The respondents were told that their participation was purely voluntary and that their rights to not respond at all were respected and their confidentiality and privacy were assured by arranging a suitable place for the interview and ensuring data were accessible to only the principal researcher.

## Results

### Socio-demographic characteristics of study participants

Out of 419 planned, 417 participated with a response rate of 99.5%. The mean age of the respondents was 28.62 years with a +5.69 standard deviation (SD). Among the respondents, 395 (94.7%) were married and 223 (53.5%) were Muslim. Concerning educational status, 216 (51.8%) of the respondents had completed primary school. The majority (76%) of the respondents were rural residents (Table 1).

**Table 1:**
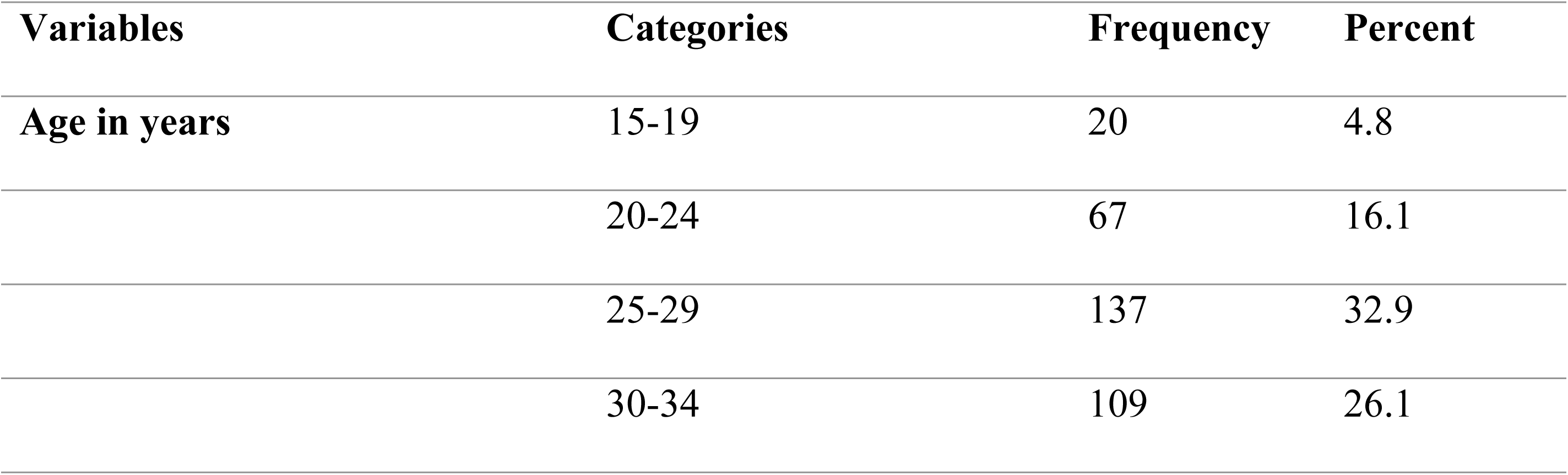
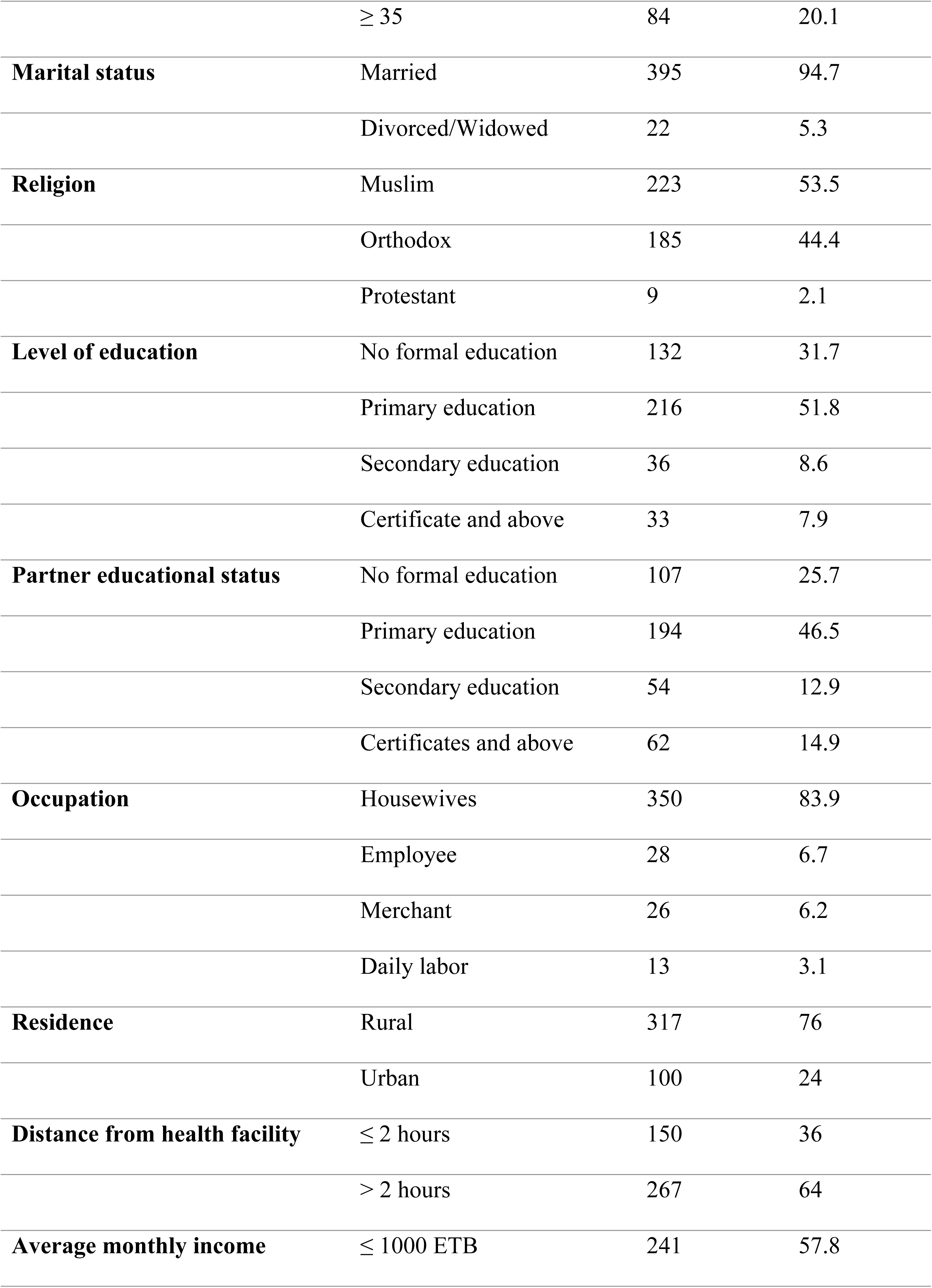
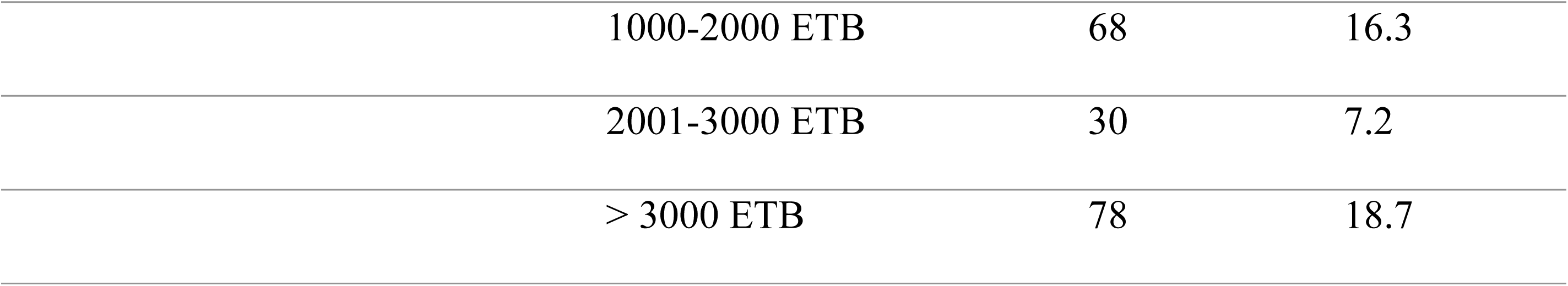
Socio-demographic characteristics of the postpartum women in Sude woreda, Arsi Zone, Oromia, Ethiopia, 2023 (n=417)

### Reproductive and reproductive health service-related characteristics

The majority (83.2%) of the respondents were multigravida and most (91.4%) of the birth was attended by midwives. The majority (89.4%) of them heard of methods of modern contraceptives and 81.8% had an intention to use modern contraceptives in the future (**Table 2**).

**Table 2:** Reproductive health service-related history of postpartum women in Sude woreda, Arsi Zone, Southeast, Ethiopia, 2023 (n=417)

| Variables | Categories | Frequency | Percent |
| --- | --- | --- | --- |
| Gravidity | Primigravida | 70 | 16.8 |
|  | Multigravida | 347 | 83.2 |
| Parity | 1 | 81 | 19.4 |
|  | 2 - 4 | 218 | 52.3 |
|  | > 4 | 118 | 28.3 |
| Number of children | ≤ 4 | 262 | 62.8 |
|  | > 4 | 155 | 37.2 |
| **The birth was attended by** | Midwives | 381 | 91.4 |
|  | Doctors | 36 | 8.6 |
| **Postpartum period** | ≤ 6 weeks | 47 | 11.3 |
|  | 7-24 weeks | 215 | 51.6 |
|  | 6-12 months | 155 | 37.2 |
| **Sexual intercourse resumed** | Yes | 257 | 61.6 |
|  | No | 160 | 38.4 |
| **Menses returned** | Yes | 232 | 55.6 |
|  | No | 185 | 44.4 |
| **Discussed PFP with the partner** | Yes | 193 | 46.3 |
|  | No | 224 | 53.7 |
| **Currently using modern contraceptives** | Yes | 240 | 57.6 |
|  | No | 177 | 42.4 |
| **If yes, what method of contraceptive using** | Injectables | 122 | 50.8 |
|  | Implants | 94 | 39.2 |
|  | Pills | 24 | 10 |
| **Heard methods of modern contraceptives** | Yes | 373 | 89.4 |
|  | No | 44 | 10.6 |
| **Intention to use contraceptives in future** | Yes | 341 | 81.8 |
|  | No | 76 | 18.2 |

### Postpartum modern contraceptive utilization

From the total of 417 respondents, 57.6% [95% CI: 52.9, 62.4)] utilized modern contraceptives. The most widely used type of modern contraceptive method was injectables (50.8%), followed by implants (39.2%) and pills (10%).

### Factors associated with postpartum modern contraceptive use

In bivariable logistic regression analysis, age, marital status, educational status, distance from the health facility, average monthly income in Ethiopian birr (ETB), gravidity, parity, number of children, discussing postpartum family planning (PPFP) with a partner, postpartum period, resumed menses, heard modern contraceptives methods, and intention to use contraceptives in the future were significantly associated with post-partum modern contraceptive (PPMC) utilization at a p-value < 0.25. After controlling the confounding effects of other variables, number of children, distance from the health facility, postpartum period, resumed menses, parity, and discussing PPMC with a partner showed a significant association with postpartum modern contraceptive utilization at a p-value < 0.05.

Accordingly, the odds of using postpartum modern contraceptives were two times higher [AOR = 2.02, 95% CI: (1.09, 3.77)] among women who had > 4 children as compared to those women who had ≤ 4 children. The odds of using postpartum modern contraceptives were more than two times higher [AOR = 2.54, 95% CI: (1.49, 4.34)] among women who were ≤ 2 hours away from the health facility compared to those women who were > 2 hours far from the health facility. The odds of using postpartum modern contraceptives were more than three times higher [AOR = 3.41, 95% CI: (1.89, 6.15)] among women between 6 to 12 months of postpartum period compared to women in the 7 to 24 weeks of postpartum period. The odds of using postpartum modern contraceptives were more than eight times higher [AOR = 8.24, 95% CI: (4.64, 14.64)] among women whose menses resumed compared to their counterparts. The odds of using postpartum modern contraceptives were three times higher [AOR = 3.10, 95% CI: (1.53, 6.29)] among women who were para 2-4 compared to those women who were greater than para 4. The odds of using postpartum modern contraceptives were nearly three times higher [AOR = 2.96, 95% CI: (1.72, 5.09)] among women who had discussed postpartum family planning (PPFP) with a partner compared to their counterparts (**Table 3**).

**Table 3:** Factors associated with modern contraceptive utilization among postpartum women in Sude woreda, Arsi Zone, Oromia, Ethiopia, 2023.

| Variables | Postpartum contraceptive use |  | COR (95% CI) | AOR (95% CI) | P-value |
| --- | --- | --- | --- | --- | --- |
|  | Yes | No |  |  |  |
| Age in years |  |  |  |  |  |
| 15-19 | 4 | 16 | 8.92 (2.72, 29.31) | 3.30 (0.52, 21.00) | 0.206 |
| 20-24 | 18 | 49 | 6.07 (2.98, 12.37) | 2.82 (0.91, 8.72) | 0.071 |
| 25-29 | 85 | 52 | 1.37 (0.77, 2.43) | 1.27 (0.58, 2.75) | 0.554 |
| 30-34 | 75 | 34 | 1.01 ( 0.55, 1.87) | 1.48 (0.70, 3.12) | 0.310 |
| ≥ 35 | 58 | 26 | 1.00 | 1.00 |  |
| Marital status |  |  |  |  |  |
| Married | 224 | 171 | 2.04 (0.78, 5.31) | 2.73 (0.70, 10.69) | 0.148 |
| Divorced | 16 | 6 | 1.00 | 1.00 |  |
| Number of children |  |  |  |  |  |
| ≤ 4 | 139 | 123 | 1.00 | 1.00 |  |
| > 4 | 101 | 54 | 0.60 (0.40, 0.91) | **2.02 (1.09, 3.77)** | 0.027 |
| Educational status |  |  |  |  |  |
| No formal Education | 82 | 43 | 1.00 | 1.00 |  |
| Primary education | 114 | 96 | 1.61 (1.02, 2.54) | 0.88 (0.46, 1.69) | 0.706 |
| Secondary education | 24 | 25 | 1.99 (1.02, 3.89) | 1.70 (0.65, 4.48) | 0.283 |
| Certificate and above | 20 | 13 | 1.24 (0.56, 2.73) | 1.18 (0.37, 3.78) | 0.784 |
| **Distance from the health facility** |  |  |  |  |  |
| ≤ 2 hours | 72 | 78 | 1.84 (1.23, 2.76) | **2.54 (1.49, 4.34)** | 0.001 |
| > 2 hours | 168 | 99 | 1.00 | 1.00 |  |
| **Average monthly income in ETB** |  |  |  |  |  |
| ≤ 1000 | 130 | 111 | 1.00 | 1.00 |  |
| 1000-2000 | 38 | 30 |  | 0.99 (0.49, 2.00) | 0.980 |
| 2001-3000 | 22 | 8 |  | 0.49 (0.17, 1.42) | 0.189 |
| > 3000 | 50 | 28 |  | 0.48 (0.23, 1.01) | 0.052 |
| **Postpartum period** |  |  |  |  |  |
| ≤ 6 weeks | 18 | 29 | 2.95 (1.54, 5.65) | 2.20 (0.93, 5.22) | 0.073 |
| 7-24 weeks | 139 | 76 | 1.00 | 1.00 |  |
| 6 - 12 months | 83 | 72 | 1.59 (1.04, 2.42) | **3.41 (1.89, 6.15)** | 0.001 |
| **Menses resumed** |  |  |  |  |  |
| Yes | 94 | 138 | 5.50 (3.54, 8.53) | **8.24 (4.64, 14.64)** | 0.001 |
| No | 146 | 39 | 1.00 | 1.00 |  |
| **Intention to use contraceptives in the future** |  |  |  |  |  |
| Yes | 207 | 134 | 0.50 (0.30, 0.82) | 0.81 (0.40, 1.65) | 0.568 |
| No | 33 | 43 | 1.00 | 1.00 |  |
| **Gravidity** |  |  |  |  |  |
| Primigravida | 16 | 54 | 6.15 (3.37, 11.20) | 3.17 (0.90, 11.09) | 0.072 |
| Multigravida | 224 | 123 | 1.00 | 1.00 |  |
| **Parity** |  |  |  |  |  |
| 1 | 21 | 60 | 8.02 (4.21, 15.27) | 2.97 (0.73, 12.15) | 0.130 |
| 2-4 | 132 | 86 | 1.83 (1.12, 2.99) | **3.10 (1.53, 6.29)** | 0.002 |
| > 4 | 87 | 31 | 1.00 | 1.00 |  |
| **Heard modern contraceptive methods** |  |  |  |  |  |
| Yes | 219 | 154 | 0.64 (0.34, 1.20) | 0.75 (0.31, 1.84) | 0.535 |
| No | 21 | 23 | 1.00 | 1.00 |  |
| **Discuss PFP with the partner.** |  |  |  |  |  |
| Yes | 85 | 108 | 2.85 (1.91, 4.26) | **2.96 (1.72, 5.09)** | 0.001 |
| No | 155 | 69 | 1.00 | 1.00 |  |

## Discussion

This study aimed to determine the prevalence of postpartum modern contraceptive utilization and identify factors that affect modern contraceptive utilization in the Sude District of Arsi Zone. The prevalence of postpartum modern contraceptive utilization was 57.6%. The current finding is comparable with the studies done in Nigeria (59.2%) (22), Egypt (54.5%) (23), South Gondar (54.7%) (17), Assosa zone (53.5%) (24), and Northwest Ethiopia (53%) (25). However, it was lower than the studies done in Malawi (75%) (26), Rwanda (67%) (27), Hossana (72.9%) (28), Arba Minch (64.7%) (29), and Debar Tabor (63%) (30). This might be due to the difference in the socio-demographic characteristics of the respondents, the time gap between the studies, the difference in the study design, and the difference in the definition of postpartum family planning (PPFP). On the other hand, the finding was higher than studies that were done in Mexico (47%) (31), India (49%) (32), Nepal (32.8%) (33), Burie District, Amhara region (20.7%) (34), Debre Berhan (41.6%) (35), Kiramu District, Western Ethiopia (28.3%) (36), and Addis Ababa (12.9%) (37). The difference might be due to the differences in socio-demographic characteristics of the study population, study design, and period.

In this study, the most common method of modern contraception used by postpartum mothers was found to be injectable (50.8%). This finding is supported by a study done in the Assosa zone (24). However, the study conducted in Arba Minch indicates that the majority of the study participants had used implants as a modern contraception method, while only 13.2% had utilized injectables (38). This disparity in results may result from variations in the methods of family planning that were available, the length of the study, and the degree of knowledge about the various family planning choices.

The findings of this study showed an association between a woman’s number of living children and her likelihood of utilizing postpartum modern contraceptives. Accordingly, the odds of using postpartum modern contraceptives were two times higher among mothers who had > 4 children as compared to those mothers who had ≤ 4 children. This finding is supported by the studies conducted in Uganda and Rwanda (27, 39). The possible justification might be having more children overall was linked to a higher likelihood of utilizing contraception after giving birth. The correlation between the total number of children and a higher likelihood of using contraception is an intriguing finding.

The current study revealed that distance from the health facility was significantly associated with postpartum modern contraceptive utilization. The odds of using postpartum modern contraceptives were more than two times higher among mothers who were ≤ 2 hours away from the health facility as compared to those mothers who were > 2 hours away from the health facility. The current finding is supported by a study conducted in Tanzania (40). The possible explanation might be utilizing postpartum modern contraceptives was higher among women who were exposed to family planning-related media than among those who were not. The possible explanation might be related to accessibility; women who are not far away from the health facility have better access to maternal health services including contraceptives compared to those who are more far away from the health facility (41).

In this study, discussing modern contraceptive methods with a partner was significantly associated with postpartum modern contraceptive use. The odds of using postpartum modern contraceptives were nearly three times higher among mothers who had discussed postpartum family planning with a partner compared to their counterparts This finding is supported by findings from Northwest Ethiopia (42), Gondar (43), Aksum (44), Burie (34), and Aroressa (45). This can be explained by the fact that any factor that influences the partner’s attitude toward contraceptives would also affect women’s use of postpartum contraceptives, either positively or negatively. Decisions about the applicability and relevance of modern contraception can be made together, increasing the likelihood that women who discuss it with their partners will be accepted and supported.

Duration of the postpartum period showed a significant association with contraceptive use. Those women between 6 to 12 months of the postpartum period had higher odds of using contraceptives when compared to women in the 7 to 24 weeks postpartum period. This finding is supported by the studies done in Nairobi (46), and Gondar town (47). This result may be explained by the fact that most women resumed menstruation after six months. Another reason might be that most women were exclusively breastfeeding for the first six months of the postpartum period.

The odds of using postpartum modern contraceptives were more than eight times higher among mothers whose menses resumed compared to their counterparts. Because they think that amenorrhea will prevent conception regardless of the length of the postpartum period, women with amenorrhea may underestimate their risk of getting pregnant. Mothers may be motivated to utilize modern contraceptives because they believe that the risk of pregnancy increases once menstruation resumes. This finding was supported by the studies done in Tanzania, South Gondar, Debre Tabor, Arba Minch, and Hossana (17, 28–30, 40).

Eventually, our results indicate that a woman’s parity was associated with her likelihood of using modern contraceptives, as grand multiparous women were much more likely to utilize modern contraceptives. Similar findings were reported in the Ethiopian Demographic and Health Survey (48) and Northwest Ethiopia (49).

## Limitations of the study

The current study has some limitations which will be addressed by future researchers. The findings may not represent the general postpartum women due to the study’s design.

## Conclusion

The postpartum modern contraceptive utilization was low compared to the previous studies. The number of children, distance from home to health facility, postpartum period, parity, and menses returning since birth, and discussing postpartum family planning with a partner were significantly associated with postpartum modern contraceptive utilization. Healthcare providers should increase postnatal care follow-up after delivery, give health information about timely contraceptive usage, and fortify the integration of family planning services with maternal and child health services. Improvements in accessibility to health facilities and encouraging postpartum family planning discussion among couples are crucial steps for enhancing modern contraceptive use among postpartum women.

## Supporting information

S1 File. English version of the information sheet and consent form. (DOCX)

S2 File. English version of the questionnaire. (DOCX)

S3 File. Dataset. (SAV)

## Acknowledgment

We would like to express our sincere gratitude and deep appreciation to the Research and Ethical Committee of Rift Valley University for the approval of ethical clearance. We would also like to extend our gratitude to the Sude Woreda Health Office for their letter of permission to conduct the study. Finally, our special thanks go to the data collectors, supervisors, and participants.

## Author Contributions

**Conceptualization:** Mohammed Sultan Gemeda, Gemechu Dereje Feyissa, Daniel G/tsadik Endashaw Mandefro.

**Data curation:** Mohammed Sultan Gemeda, Gemechu Dereje Feyissa, Daniel G/tsadik Endashaw Mandefro.

**Formal analysis:** Mohammed Sultan Gemeda, Gemechu Dereje Feyissa.

**Investigation:** Mohammed Sultan Gemeda.

**Methodology:** Mohammed Sultan Gemeda, Gemechu Dereje Feyissa, Daniel G/tsadik, Endashaw Mandefro.

**Project administration:** Mohammed Sultan Gemeda, Gemechu Dereje Feyissa.

**Resources:** Mohammed Sultan Gemeda, Gemechu Dereje Feyissa, Mokonnen Dereje.

**Software:** Mohammed Sultan Gemeda, Gemechu Dereje Feyissa, Mokonnen Dereje.

**Supervision:** Mohammed Sultan Gemeda, Daniel G/tsadik, Endashaw Mandefro, Hunde Lemi.

**Validation:** Mohammed Sultan Gemeda, Gemechu Dereje Feyissa, Endashaw Mandefro.

**Visualization:** Mohammed Sultan Gemeda, Daniel G/tsadik, Hunde Lemi, Mokonnen Dereje.

**Writing – original draft:** Mohammed Sultan Gemeda, Gemechu Dereje Feyissa.

**Writing – review & editing:** Mohammed Sultan Gemeda, Gemechu Dereje Feyissa, Hunde Lemi.

## Data availability statement

All relevant data are within the manuscript and its supporting information files.

## Funding

Not applicable.

## Consent for publication

Not applicable.

## Competing interests

The authors declare that they have no competing interests.

## Abbreviations

AOR: Adjusted Odd Ratio
CI: Confidence Interval
COR: Crude Odds Ratio
EDHS: Ethiopian Demographic Health Survey
FP: Family Planning
PPFP: Postpartum Family Planning
SPSS: Statistical Package for Social Sciences

## Notes

### Competing Interest Statement

The authors have declared no competing interest.

### Funding Statement

The author(s) received no specific funding for this work.

